# Fine-tuning and pre-training improve the predictive accuracy of large language models for rheumatoid arthritis disease activity

**DOI:** 10.1101/2024.10.14.24315448

**Authors:** Suguru Honda, Katsunori Ikari, Mayuko Fujisaki, Eiichi Tanaka, Masayoshi Harigai

**Author notes:** **Corresponding author:** Masayoshi Harigai, MD, PhD, Department of Rheumatology, Sanno Hospital, Tokyo, Japan, 8-10-16 Akasaka, Minato-ku, Tokyo, 107-0052, Japan.

## Abstract

**Objective:** To evaluate whether the performance of the large language model (LLM) Llama2 improves with pre-training and fine-tuning, and to compare its predictive accuracy with that of a linear regression model for rheumatoid arthritis (RA) disease activity.

**Methods:** Clinical data from 11,865 patients in the cohort were used to predict disease activity at two years on four indices (Disease Activity Score (DAS) 28-Erythrocyte sedimentation rate (ESR), DAS28-C-reactive protein (CRP), Clinical Disease Activity Index (CDAI) or Japanese Health Assessment Questionnaire (J-HAQ)). Logistic regression was employed for the linear model for comparison. The predictive performance was assessed using area under the curve (AUC). Additional performance metrics including precision, recall, and F1 score were calculated.

**Results:** Pre-training significantly improved AUC of Meditron (Llama2 pre-trained with medical data) for DAS28-ESR >5.1, DAS28-ESR <2.6, DAS28-CRP <2.3, J-HAQ score >2.5, and J-HAQ score <0.5 (*P* <0.05). Fine-tuning resulted in significant improvements in AUC for Llama2 across all indices (*P* <0.05) except CDAI >22, and for Meditron in DAS28-ESR <2.6, DAS28-CRP >4.1, DAS28-CRP <2.3 and CDAI ≤2.8 (*P* <0.05). Both LLMs significantly outperformed linear regression in predicting DAS28-ESR <2.6, DAS28-CRP >4.1, DAS28-CRP <2.3, J-HAQ score >2.5, and J-HAQ score <0.5 (*P* <0.05). Furthermore, DAS28-CRP >4.1, DAS28-CRP <2.3, J-HAQ score >2.5 and J-HAQ score <0.5, Llama2 or Meditron consistently outperformed linear regression across all performance metrics.

**Conclusion:** Both pre-training and fine-tuning significantly improved the performance of Llama2. Both LLMs outperformed the linear regression model in predicting 5 out of the 8 categories of indices.

## INTRODUCTION

In the past few years, advancements in Large Language Models (LLMs) have shown significant potential for improving the management and diagnosis of diseases, extracting clinical information from Electronic Medical Records (EMRs), and assisting in the creation of pathology and imaging reports (1–3). Literature increasingly recognizes the utility of LLMs as clinical decision support tools that could aid in clinical diagnostics. The prospective integration of LLMs into EMR systems is anticipated to enhance aspects of medical practice, research, and education, although there are several critical challenges to put this into practice.

Accuracy remains a crucial concern. For instance, while models like GPT-4 have demonstrated capabilities that surpass the thresholds of medical board examinations in diagnostic and treatment knowledge domains (4), their application in predicting disease activity indices—which are essential for managing conditions like rheumatoid arthritis (RA)—has not been adequately addressed. These indices are vital for physicians to adjust treatment plans based on disease activity. Therefore, assessing the ability of LLMs to predict disease activity with high precision is essential for their future roles in clinical support systems.

Additionally, security and privacy issues pose substantial challenges. The use of cloud-based models like GPT-4, which operate with non-transparent sources and processes, raises concerns regarding data breaches and unauthorized access (5,6). In contrast, open-source models like Llama2 (7), which can be deployed locally, offer advantages in data security as they allow for internal data processing. However, these models often do not provide the same level of accuracy in medical applications as cloud-based models.

Recent initiatives have aimed to enhance the medical accuracy of open-source models through supplementary pre-training with medical datasets. The Meditron model, an adaptation of Llama2 further trained with clinical practice guidelines and literature, exemplifies this approach, improving its accuracy on medical reasoning benchmarks such as MedQA from 63.8% to 70.2% (8). Moreover, fine-tuning has proven effective in adapting these models for specific medical tasks, yielding outcomes that sometimes surpass those of medical experts, particularly in the domain of medical text summarization (1).

Despite these technological advances, the literature still lacks comprehensive studies on the reliability and precision of pre-trained and fine-tuned model responses, especially in the field of rheumatology. Although several studies have examined model responses without pre-training or fine-tuning (9–11), to our knowledge, no studies have examined the accuracy of pre-trained and fine-tuned LLM models for disease activity in RA. In this study, we aim to develop a secure, high-accuracy, open-source model for predicting disease activity in patients with RA by utilizing pre-training and fine-tuning and comparing its performance with a linear regression model.

## MATERIALS AND METHODS

### Study population

Clinical data were obtained from the Institute of Rheumatology, Rheumatoid Arthritis (IORRA) cohort, which was initiated in 2000. The IORRA cohort is a large, single-center observational study of Japanese patients with RA, comprising approximately 0.5% of the total Japanese RA population (12). From 2000 to 2022, a total of 15,004 patients were enrolled. For this study, we selected cases in which at least one disease activity index, including Disease Activity Score (DAS) 28-Erythrocyte sedimentation rate (ESR), DAS28-C-reactive protein (CRP), Clinical Disease Activity Index (CDAI) or Japanese Health Assessment Questionnaire (J-HAQ) score, had been followed for at least two years (n = 11,865) (Figure 1). To prevent data leakage during model training, we first selected 102 cases for the validation set and 2,263 cases for the test set. The remaining cases were used as the training set (n = 9,500).

**Figure 1.**
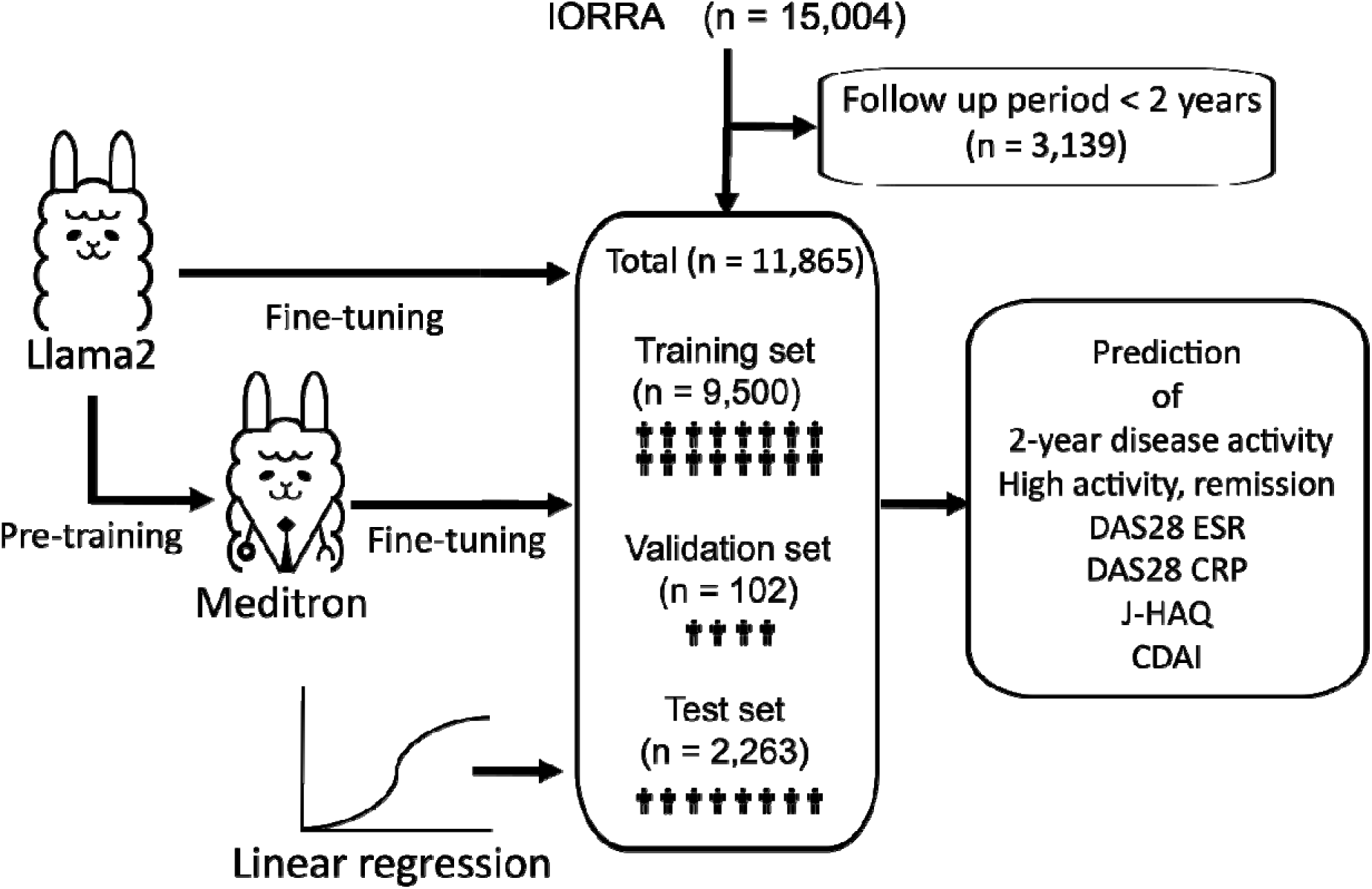
Overview of study design and workflow. The IORRA cohort (n = 15,004) was used for this study, with patients who had a follow-up period of less than two years excluded (n = 3,139). The remaining data were divided into three sets: a training set (n = 9,500), a validation set (n = 102), and a test set (n = 2,263). Llama2 and Meditron served as the large language models (LLMs) for predicting disease activity and physical function. Meditron, pre-trained on medical literature, were fine-tuned using the data of training set. A linear regression was used for comparison. Predictive indicators were DAS28-ESR, DAS28-CRP, and CDAI (high disease activity or remission), and J-HAQ score (high score or remission) after two years.

### Data conversion

The extracted data from the IORRA cohort were converted into text-based problem statements using in-house scripts. For example, if a patient with ID1 had the following characteristics: Age 72, Sex 1, Methotrexate 1, Prednisone 0, and Hypertension 1, the corresponding text would be:

*"A 72-year-old female rheumatoid arthritis patient treated with methotrexate … The patient has a history of hypertension. What is the probability that this patient’s CDAI will be remission at 2 years?"*

These text-based questions, paired with the respective answers, were stored in JSON format to create the training dataset.

### Model Selection

For this study, we utilized two LLMs: Llama2 (70B) and Meditron (70B). Both models are autoregressive and open source, which allowed for local deployment, ensuring data privacy and security during model development. Llama2, developed by Meta, is a widely used open-source model, known for its instruction-following capabilities and extended context lengths (7). We selected the 70B version of Llama2 to benefit from its larger parameter size, which provides improved performance on complex tasks.

The Meditron is based on Llama2 and further pretrained on a large corpus of medical literature, including PubMed articles, abstracts, and internationally recognized clinical practice guidelines (8). This additional medical domain-specific pretraining allows Meditron to outperform standard LLMs like GPT-3.5 on medical reasoning tasks. Meditron has demonstrated competitive accuracy on benchmarks such as MedQA and PubMedQA, making it a suitable candidate for the prediction of disease activity in patients with RA

### Fine-tuning

To fine-tune the Llama2 and Meditron (70B) models for the prediction of disease activity in patients with RA, we employed the QLoRA fine-tuning technique. QLoRA is an efficient, low-rank adaptation method that allows fine-tuning with minimal changes to the original model weights by inserting trainable matrices into the attention layers (13). This method was chosen to minimize computational resources while maintaining the integrity of the base model.

The fine-tuning was conducted with a learning rate of 3e-5, a batch size of 2, and gradient accumulation steps of 8. LoRA parameters included a rank of 8 and an alpha of 16. Based on the dataset and batch size, the number of epochs was approximately 1 epoch.

### Statistical analysis

DAS28-ESR, DAS28-CRP, CDAI were categorized as high disease activity or remission and J-HAQ score was categorized as high score or remission two years after the baseline assessment (Supplemental Table 1). The explanatory variables used in the model included demographics, disease activity, laboratory data, medication use, and medical history (details are provided in Supplemental Table 2). For comparison of event prediction accuracy between the LLMs and traditional statistical models, we used a logistic regression model as the linear baseline. We primarily used the AUC from receiver operating characteristic (ROC) analysis to assess model performance. To compare the AUCs between models, we applied DeLong’s test. Values of *P□*<□0.05 were considered significant. No corrections were made for multiple comparisons. Other metrics, including sensitivity, specificity, positive predictive value (PPV), negative predictive value (NPV), precision (the proportion of true positive predictions among all positive predictions made by the model), recall (the proportion of true positive predictions among all actual positive cases), and F1 score (the harmonic mean of precision and recall, providing a balance between the two), were calculated using Python’s sklearn library. The development environment was Python 3.8.16 running on two parallel NVIDIA RTX 3090 24 GB GPUs and a 6000 Ada 48GB GPU for fine-tuning the LLM.

## RESULTS

### Patients’ characteristics

The IORRA cohort in this study includes 11,865 patients with RA, predominantly female (82.9%), with a mean age of 63.0 years and an average disease duration of 15.4 years. A significant portion (33.6%) have a history of smoking (see Supplemental Table 2). The average DAS28-ESR is 2.9, DAS28-CRP is 2.3, and the CDAI is 6.0, all of which met the criteria for low disease activity. Methotrexate is the most used conventional synthetic disease-modifying antirheumatic drugs (csDMARD) (64.1%), and 40.8% of patients are on prednisolone. The cohort is characterized by older patients with long-standing RA, many of whom are on csDMARDs or biologic DMARDs.

### Impact of Pre-training and Fine-Tuning on Model Performance

First, to determine whether Meditron, the model in which Llama2 was pre-trained with medical knowledge, had acquired accurate knowledge of disease activity in RA, both models were asked the following question about the DAS28-ESR: “*How is the DAS28-ESR calculated in patients with rheumatoid arthritis? First, what variables are needed to calculate the DAS28-ESR? Second, please provide and explain the calculation formula. Finally, please give us your interpretation of the DAS28-ESR.*” The answer of Llama2 was incorrect, with the formula “DAS28-ESR = 0.56 × √(SJC + TJC) + 0.28 × PGA + 0.11 × ESR,” while Meditron provided the correct formula, “DAS28-ESR = 0.56 × √(tender28) + 0.28 × √(swollen28) + 0.70 × ln(esr) + 0.014 × global." Furthermore, Meditron accurately defined the criteria for remission and high disease activity, whereas the explanation of Llama2 was inaccurate (see Supplemental Table 3).

The effect of pre-training on predicting the categories of disease activity or J-HAQ score was assessed by comparing the AUC of Meditron and Llama2 without fine-tuning using several indices. Meditron without fine-tuning (red line in Figure 2) significantly outperformed Llama2 without fine-tuning (blue line in Figure 2) for DAS28-ESR >5.1, DAS28-ESR <2.6, DAS28-CRP <2.3, J-HAQ score >2.5, and J-HAQ score <0.5 (p < 0.05, see Supplemental Table 4).

**Figure 2.**
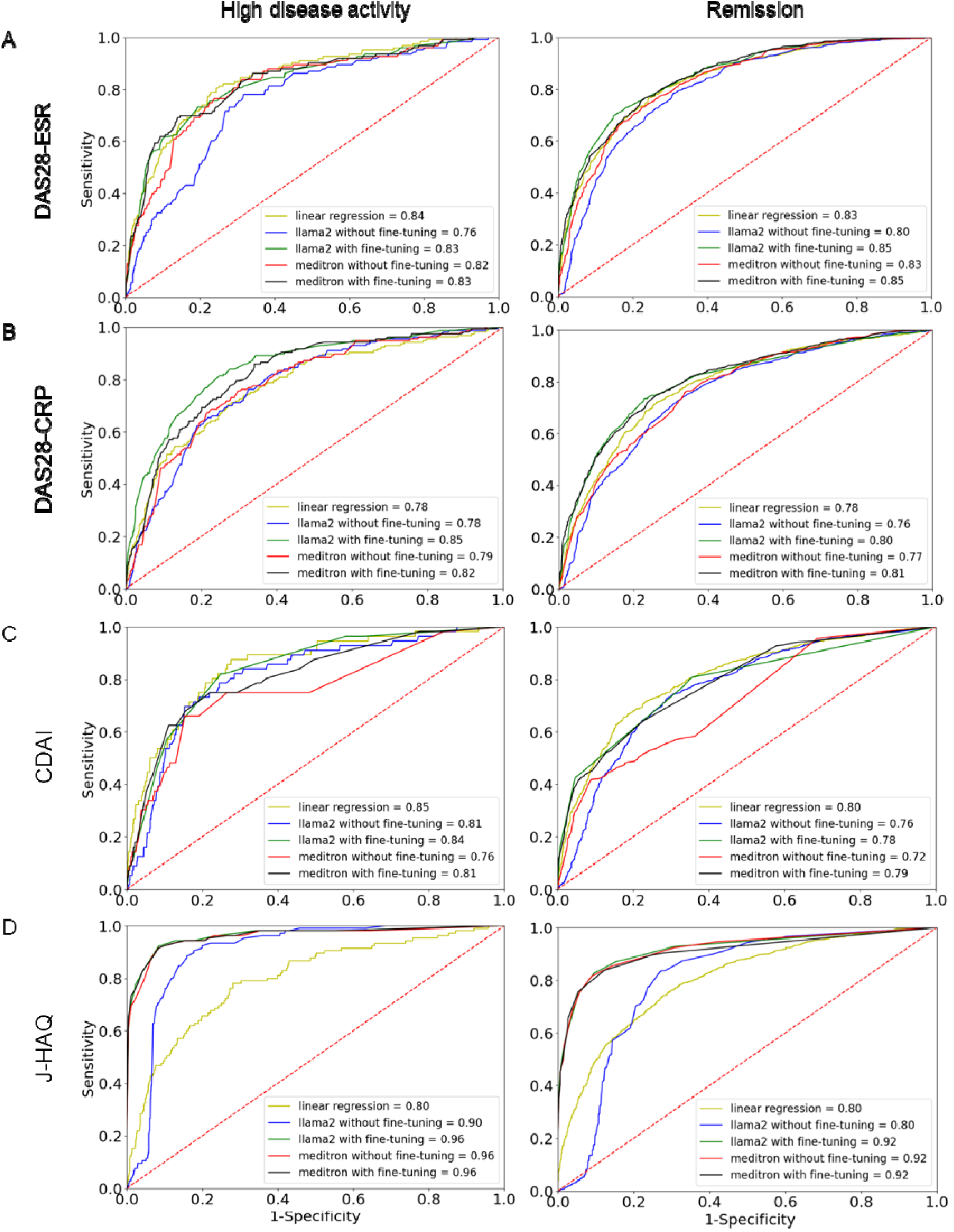
Comparison of predictive performance between models across multiple indices. Receiver operating characteristic (ROC) curves displaying the classification performance of five models in predicting high disease activity and remission across three disease activity indices and high score and remission in J-HAQ in patients with rheumatoid arthritis: DAS28-ESR (A), DAS28-CRP (B), CDAI (C), and J-HAQ (D). The models evaluated include a linear regression model, Llama2 without fine-tuning, Llama2 with fine-tuning, Meditron without fine-tuning, and Meditron with fine-tuning. For DAS28-ESR (A), the ROC curves on the left show the models’ performance in predicting high disease activity (DAS28-ESR >5.1), while the curves on the right assess the models’ performance in predicting remission (DAS28-ESR <2.6). A similar layout is followed for DAS28-CRP (>4.1 for high disease activity and <2.3 for remission) (B), and CDAI (>22 for high disease activity and ≤2.8 for remission) (C). For J-HAQ (D), the left and right panel show the models’ performance in predicting J-HAQ score >2.5 and J-HAQ <0.5, respectively.

Similarly, the effect of fine-tuning was assessed in Llama2 and Meditron models, separately. Fine-tuning of Llama2 (green line vs. blue line in Figure 2) resulted in a significant improvement in AUC values across all indices (p < 0.05, see Supplemental Table 5), except for the prediction of CDAI >22 (P = 0.35). For Meditron (black line vs. red line in Figure 2), fine-tuning led to significant improvements in AUC for DAS28 ESR <2.6, DAS28 CRP >4.1, DAS28 CRP <2.3 and CDAI ≤2.8 (p < 0.05, see supplemental Table 5).

### Comparison of AUC Between Linear Regression and Fine-tuned LLM Models

The performance of fine-tuned Llama2 was compared to that of a linear regression model using ROC analysis for the four indices (Figure 2, Supplemental Table 6). Fine-tuned Llama2 (green line vs. yellow line in Figure 2) significantly outperformed the linear regression model in most indices, with marked improvements in predicting DAS28-ESR <2.6 (P = 0.0042, right panel in Figure 2A), DAS28-CRP >4.1 (P < 0.0001, left panel in Figure 2B), DAS28 CRP <2.3 (P = 0.020, right panel in Figure 2B), J-HAQ >2.5 (P < 0.0001, left panel in Figure 2D), and J-HAQ <0.5 (P < 0.0001, right panel in Figure 2D). However, there was no significant difference in AUC for predicting DAS28-ESR >5.1 (P = 0.44, left panel in Figure 2A) and CDAI >22 (P = 0.58, left panel in Figure 2C). The linear regression model showed a significant advantage in predicting CDAI ≤2.8 (P = 0.0082, right panel in Figure 2C).

For Meditron, the fine-tuned model (black line vs. yellow line in Figure 2) was also compared to the linear regression model (Supplemental Table 6). Meditron significantly outperformed the linear regression model across most indices, with substantial improvements in DAS28-ESR <2.6 (P = 0.012, right panel in Figure 2A), DAS28-CRP >4.1 (P = 0.0087, left panel in Figure 2B), DAS28-CRP <2.3 (P = 0.0010, left panel in Figure 2B), J-HAQ score >2.5 (P < 0.0001, left panel in Figure 2D), and J-HAQ <0.5 (P < 0.0001, right panel in Figure 2D). However, no significant differences were observed for DAS28-ESR >5.1 (P = 0.54, left panel in Figure 2A), CDAI >22 (P = 0.14, left panel in Figure 2C), and CDAI ≤2.8 (P = 0.058, right panel in Figure 2C).

### Performance Metrics Comparison Across Linear Regression, Llama2, and Meditron Models

The performance metrics for linear regression, Llama2, and Meditron models were compared across multiple disease activity indices, including sensitivity, specificity, PPV, NPV, precision, recall, and F1 score (Table 1). Notably, for DAS28-CRP >4.1, DAS28-CRP <2.3, J-HAQ score >2.5, and J-HAQ score <0.5, LLMs – either Llama2 or Meditron – outperformed the linear regression model across all metrics. Furthermore, for Specificity and F1 score, Llama2 or Meditron consistently achieved higher values compared to the linear regression model across these indices. For example, Meditron demonstrated superior specificity in DAS28-ESR >5.1 (Specificity = 0.85) and J-HAQ score >2.5 (Specificity = 0.92), while Llama2 achieved the highest F1 score for DAS28-CRP >4.1, DAS28-CRP <2.1, J-HAQ score >2.5, J-HAQ score <0.5, and CDAI ≤2.8 (F1 = 0.32, 0.70, 0.49, 0.85, and 0.78, respectively).

**Table 1.**
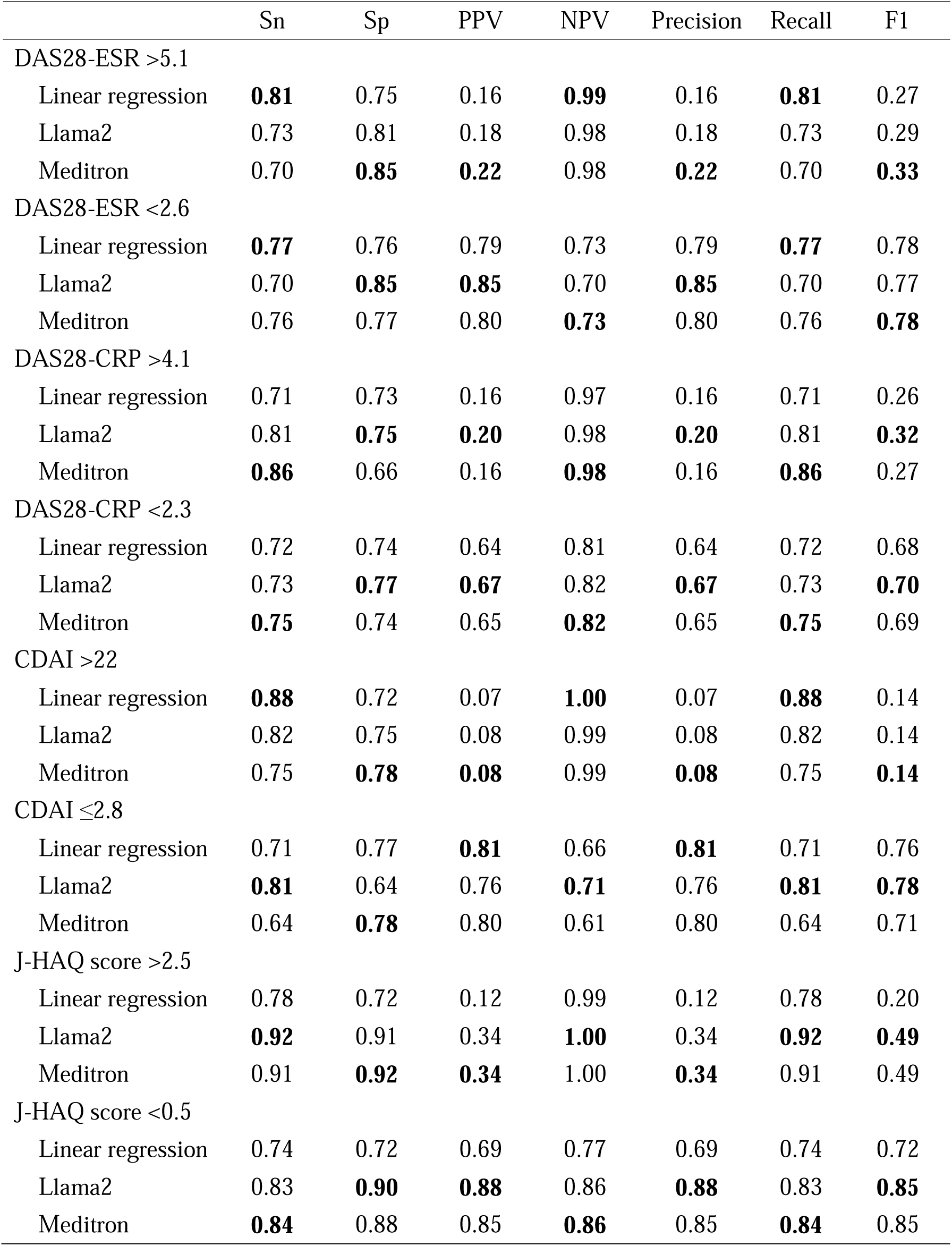
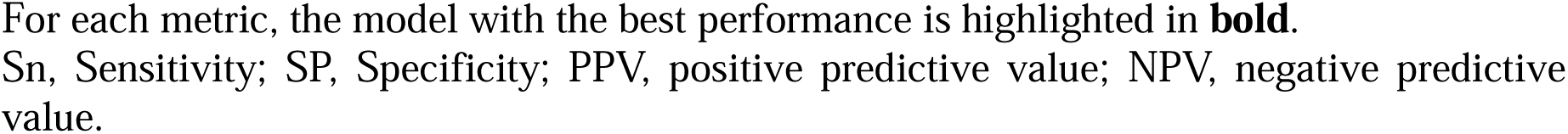
Comparison of performance metrics between linear regression, Llama2, and Meditron across multiple disease activity indices.

|  | Sn | Sp | PPV | NPV | Precision | Recall | F1 |
| --- | --- | --- | --- | --- | --- | --- | --- |
| DAS28-ESR >5.1 |  |  |  |  |  |  |  |
| Linear regression | **0.81** | 0.75 | 0.16 | **0.99** | 0.16 | **0.81** | 0.27 |
| Llama2 | 0.73 | 0.81 | 0.18 | 0.98 | 0.18 | 0.73 | 0.29 |
| Meditron | 0.70 | **0.85** | **0.22** | 0.98 | **0.22** | 0.70 | **0.33** |
| DAS28-ESR <2.6 |  |  |  |  |  |  |  |
| Linear regression | **0.77** | 0.76 | 0.79 | 0.73 | 0.79 | **0.77** | 0.78 |
| Llama2 | 0.70 | **0.85** | **0.85** | 0.70 | **0.85** | 0.70 | 0.77 |
| Meditron | 0.76 | 0.77 | 0.80 | **0.73** | 0.80 | 0.76 | **0.78** |
| DAS28-CRP >4.1 |  |  |  |  |  |  |  |
| Linear regression | 0.71 | 0.73 | 0.16 | 0.97 | 0.16 | 0.71 | 0.26 |
| Llama2 | 0.81 | **0.75** | **0.20** | 0.98 | **0.20** | 0.81 | **0.32** |
| Meditron | **0.86** | 0.66 | 0.16 | **0.98** | 0.16 | **0.86** | 0.27 |
| DAS28-CRP <2.3 |  |  |  |  |  |  |  |
| Linear regression | 0.72 | 0.74 | 0.64 | 0.81 | 0.64 | 0.72 | 0.68 |
| Llama2 | 0.73 | **0.77** | **0.67** | 0.82 | **0.67** | 0.73 | **0.70** |
| Meditron | **0.75** | 0.74 | 0.65 | **0.82** | 0.65 | **0.75** | 0.69 |
| CDAI >22 |  |  |  |  |  |  |  |
| Linear regression | **0.88** | 0.72 | 0.07 | **1.00** | 0.07 | **0.88** | 0.14 |
| Llama2 | 0.82 | 0.75 | 0.08 | 0.99 | 0.08 | 0.82 | 0.14 |
| Meditron | 0.75 | **0.78** | **0.08** | 0.99 | **0.08** | 0.75 | **0.14** |
| CDAI ≤2.8 |  |  |  |  |  |  |  |
| Linear regression | 0.71 | 0.77 | **0.81** | 0.66 | **0.81** | 0.71 | 0.76 |
| Llama2 | **0.81** | 0.64 | 0.76 | **0.71** | 0.76 | **0.81** | **0.78** |
| Meditron | 0.64 | **0.78** | 0.80 | 0.61 | 0.80 | 0.64 | 0.71 |
| J-HAQ score >2.5 |  |  |  |  |  |  |  |
| Linear regression | 0.78 | 0.72 | 0.12 | 0.99 | 0.12 | 0.78 | 0.20 |
| Llama2 | **0.92** | 0.91 | 0.34 | **1.00** | 0.34 | **0.92** | **0.49** |
| Meditron | 0.91 | **0.92** | **0.34** | 1.00 | **0.34** | 0.91 | 0.49 |
| J-HAQ score <0.5 |  |  |  |  |  |  |  |
| Linear regression | 0.74 | 0.72 | 0.69 | 0.77 | 0.69 | 0.74 | 0.72 |
| Llama2 | 0.83 | **0.90** | **0.88** | 0.86 | **0.88** | 0.83 | **0.85** |
| Meditron | **0.84** | 0.88 | 0.85 | **0.86** | 0.85 | **0.84** | 0.85 |

For each metric, the model with the best performance is highlighted in **bold**.
Sn, Sensitivity; SP, Specificity; PPV, positive predictive value; NPV, negative predictive value.

## DISCUSSION

This study represents the first attempt to compare the performance of pre-trained and fine-tuned LLMs with that of a linear regression model in predicting categories of disease activity and physical function in patients with RA. Our findings demonstrate that both pre-training and fine-tuning significantly enhanced the predictive accuracy of Llama2 in most cases. Furthermore, both LLMs outperformed a linear regression model in predicting DAS28-ESR <2.6, DAS28-CRP >4.1 and DAS28-CRP <2.3, J-HAQ scrore >2.5, and J-HAQ score <0.5, while maintaining consistent superiority across multiple performance metrics. These results lay the foundation for constructing robust LLMs that not only ensure high predictive accuracy but also can be securely integrated into local EMR systems, offering a promising tool to support clinical decision-making.

Even without fine-tuning, pre-training alone significantly improved the performance of Meditron compared to Llama2. This improvement can be attributed to Meditron’s pre-training on medical literature and guidelines, which enabled the model to acquire domain-specific knowledge about RA and related conditions. For instance, the model could understand complex clinical relationships, such as how IL-6 inhibitors can cause CRP levels to normalize or how comorbidities can lead to higher HAQ scores. With this foundational knowledge, Meditron could make more accurate predictions without the need for additional fine-tuning. These nuanced understandings are challenging for traditional linear models to capture, highlighting the unique capabilities of LLMs to comprehend and integrate broader clinical scenarios.

Interestingly, despite the expected advantages of pre-training followed by fine-tuning, Meditron’s fine-tuned model showed only comparable performance when compared to Llama2’s fine-tuned model. One possible explanation is the occurrence of catastrophic forgetting, a phenomenon where a model loses previously acquired knowledge during the fine-tuning process (14). To explore this, we asked the fine-tuned Meditron model the same question regarding DAS28 ESR calculation that was posed during pre-training evaluation, and it still provided a correct and accurate response (see Supplemental Table 7). This suggests that catastrophic forgetting may not be the primary issue. Instead, it is possible that Meditron’s pre-trained knowledge provided diminishing returns when applied to the structured cohort data used in this study. A previous research has shown that LLMs excel in interpreting unstructured data, such as physician notes in electronic health records (2), and their advantages may be less pronounced when working with highly structured clinical datasets, as was the case here. This may explain why the performance difference between Meditron and Llama2 were modest after fine-tuning.

This study has several limitations. First, the dataset used for training and testing was derived from a single-center cohort, which may limit the generalizability of the findings to other populations. Additionally, while we utilized a comprehensive set of disease activity indices and physical function index, the models were not tested on more complex, unstructured clinical data such as physicians’ notes or imaging data, where LLMs may demonstrate superior performance.

In conclusion, both pre-training and fine-tuning significantly enhanced LLM’s performance in predicting the categories of disease activity or physical function. Both Llama2 and Meditron outperformed the linear regression model in the key indices, highlighting their potential to support clinical decision-making. This study lays the groundwork for future applications of LLMs in secure, effective management of patients with RA.

## Contributors

M.H. and S.H. conceived the study design. S.H. conducted all of the analyses with the help of K.I., and S.H. drafted the manuscript. K.I., M.F., S.H., E.T., and M.H. collected samples and clinical information. All authors critically reviewed and approved the manuscript.

## Acknowledgment

We thank all patients in the IORRA database, and all members of the Institute of Rheumatology, Tokyo Women’s Medical University Hospital for the successful management of the IORRA study cohort. The IORRA cohort was supported by unrestricted grants from six pharmaceutical companies: AbbVie Japan GK, Asahi Kasei Pharma Co., Ayumi Pharmaceutical Co., Taisho Pharma Co., Ltd., Teijin Pharma Ltd., and Bristol-Myers Squibb Company.

## Ethics approval

The IORRA cohort study (#2952-R and #2922-R16) were approved by the ethics committee of Tokyo Women’s Medical University, and informed consent was obtained from all patients before each survey.

## Patient and public involvement

Patients and/or the public were not involved in the design, or conduct, or reporting, or dissemination plans of this research.

## Patient consent for publication

Not required.

## Data sharing statement

The data using analysis that support the findings of this study are available on reasonable request to the authors.

## Code availability

The QLoRA code is available at https://github.com/artidoro/qlora.

Our in-house script for generating the JSON file is available at https://github.com/honda-s691470/IOR_LLM.

## REFERENCES

1. Van Veen D, Van Uden C, Blankemeier L, Delbrouck JB, Aali A, Bluethgen C, et al. Adapted large language models can outperform medical experts in clinical text summarization. Nat Med. 2024 Apr;30(4):1134–42.

2. Jiang LY, Liu XC, Nejatian NP, Nasir-Moin M, Wang D, Abidin A, et al. Health system-scale language models are all-purpose prediction engines. Nature. 2023 Jul;619(7969):357–62.

3. Tu T, Azizi S, Driess D, Schaekermann M, Amin M, Chang PC, et al. Towards Generalist Biomedical AI. NEJM AI [Internet]. 2024 Feb 22 [cited 2024 Oct 5];1(3). Available from: https://ai.nejm.org/doi/10.1056/AIoa2300138

4. Nori H, King N, McKinney SM, Carignan D, Horvitz E. Capabilities of GPT-4 on Medical Challenge Problems [Internet]. arXiv; 2023 [cited 2024 Oct 5]. Available from: https://arxiv.org/abs/2303.13375

5. Venerito V, Bilgin E, Iannone F, Kiraz S. AI am a rheumatologist: a practical primer to large language models for rheumatologists. Rheumatology (Oxford). 2023 Oct 3;62(10):3256–60.

6. Benavent D, Madrid-García A. Large language models and rheumatology: are we there yet? Rheumatology Advances in Practice. 2024 Sep 18;rkae119.

7. Touvron H, Martin L, Stone K, Albert P, Almahairi A, Babaei Y, et al. Llama 2: Open Foundation and Fine-Tuned Chat Models [Internet]. arXiv; 2023 [cited 2024 Oct 5]. Available from: https://arxiv.org/abs/2307.09288

8. Chen Z, Cano AH, Romanou A, Bonnet A, Matoba K, Salvi F, et al. MEDITRON-70B: Scaling Medical Pretraining for Large Language Models [Internet]. arXiv; 2023 [cited 2024 Oct 5]. Available from: https://arxiv.org/abs/2311.16079

9. Ye C, Zweck E, Ma Z, Smith J, Katz S. Doctor Versus Artificial Intelligence: Patient and Physician Evaluation of Large Language Model Responses to Rheumatology Patient Questions in a Cross-Sectional Study. Arthritis Rheumatol. 2024 Mar;76(3):479–84.

10. Rusinovich Lovgach O, Calvo-Aranda E, Ramos-Lisbona AI, Cardoso-Peñafiel P, Navarro Palomo P, Machattou M, et al. POS0444 ARTIFICIAL INTELLIGENCE VERSUS RHEUMATOLOGIST IN DECISION MAKING IN THE TREATMENT OF RHEUMATOID ARTHRITIS. DO WE THINK ALIKE? In: Scientific Abstracts [Internet]. BMJ Publishing Group Ltd and European League Against Rheumatism; 2024 [cited 2024 Oct 6]. p. 467.1-467. Available from: https://ard.bmj.com/lookup/doi/10.1136/annrheumdis-2024-eular.1032

11. Coskun BN, Yagiz B, Ocakoglu G, Dalkilic E, Pehlivan Y. Assessing the accuracy and completeness of artificial intelligence language models in providing information on methotrexate use. Rheumatol Int. 2024 Mar;44(3):509–15.

12. Yamanaka H, Tanaka E, Nakajima A, Furuya T, Ikari K, Taniguchi A, et al. A large observational cohort study of rheumatoid arthritis, IORRA: Providing context for today’s treatment options. Mod Rheumatol. 2020 Jan;30(1):1–6.

13. Dettmers T, Pagnoni A, Holtzman A, Zettlemoyer L. QLoRA: Efficient Finetuning of Quantized LLMs [Internet]. arXiv; 2023 [cited 2024 Oct 5]. Available from: https://arxiv.org/abs/2305.14314

14. Luo Y, Yang Z, Meng F, Li Y, Zhou J, Zhang Y. An Empirical Study of Catastrophic Forgetting in Large Language Models During Continual Fine-tuning [Internet]. arXiv; 2023 [cited 2024 Oct 5]. Available from: https://arxiv.org/abs/2308.08747

