## Supplemental Table for "Fine-tuning and pre-training improve the predictive accuracy of large language models for rheumatoid arthritis disease activity"

**Table S1**. **Definitions of remission and high disease activity/high score for each index**

|  | remission | high disease activity or high score |
| --- | --- | --- |
| DAS28-ESR | <2.6 | >5.1 |
| DAS28-CRP | <2.3 | >4.1 |
| CDAI | ≤ 2.8 | >22 |
| J-HAQ | <0.5 | >2.5 |

CDAI, Clinical Disease Activity Index; CRP, C-reactive protein; DAS, Disease Activity Score; ESR, Erythrocyte Sedimentation Rate; J-HAQ, Japanese Health Assessment Questionnaire;

**Table S2. Patients’ characteristics**

|  | n = 11,865 |
| --- | --- |
| Demographics |  |
| Female, n (%) | 9836 (82.9) |
| Age, mean (SD) | 63.0 (13.9) |
| Disease duration, mean (SD) year | 15.4 (10.7) |
| BMI, mean (SD) | 21.5 (3.3) |
| Previous or current smoker, n (%) | 3987 (33.6) |
| Disease activity |  |
| DAS28-ESR, mean (SD) | 2.9 (1.2) |
| DAS28-CRP, mean (SD) | 2.3 (1.0) |
| CDAI, mean (SD) | 6.0 (6.2) |
| J-HAQ, mean (SD) | 0.7 (0.8) |
| Tender joint count 45, mean (SD) | 1.3 (3.1) |
| Swollen joint count 45, mean (SD) | 1.3 (2.5) |
| Patient visual analogue scale, mean (SD) | 25.5 (25.5) |
| Physician visual analogue scale, mean (SD) | 11.0 (13.9) |
| Laboratory data |  |
| Rheumatoid factor, mean (SD) IU/mL | 133.8 (317.4) |
| Anti-cyclic citrullinated peptide antibody, n (%) U/ml | 9539 (80.4) |
| C-reactive protein, mean (SD) mg/L | 6.3 (12.8) |
| Erythrocyte Sedimentation Rate, mean (SD) mm/hr | 27.2 (22.3) |
| Creatinine, mean (SD) mg/dl | 0.7 (0.3) |
| Hemoglobin, mean (SD) mg/dl | 12.6 (1.4) |
| White blood cell, mean (SD) uL | 6363.7 (2192.1) |
| Aspartate aminotransferase, mean (SD) U/L | 20.5 (24.3) |
| Alanine aminotransferase, mean (SD) U/L | 24.0 (15.0) |
| Total protein, mean (SD) g/dL | 7.2 (0.5) |
| Medication use |  |
| Methotrexate, n (%) | 7604 (64.1) |
| Bucillamine, n (%) | 1368 (11.5) |
| Salazosulfapyridine, n (%) | 2165 (18.2) |
| Tacrolimus, n (%) | 1034 (8.7) |
| Iguratimod, n (%) | 380 (3.2) |
| Prednisolone, n (%) | 4841 (40.8) |
| Infliximab, n (%) | 151 (12.7) |
| Etanercept, n (%) | 607 (5.1) |
| Tocilizumab, n (%) | 562 (4.7) |
| Adalimumab, n (%) | 146 (1.2) |
| Abatacept, n (%) | 214 (1.8) |
| Golimumab, n (%) | 206 (1.7) |
| Certolizumab pegol, n (%) | 80 (0.7) |
| Sarilumab, n (%) | 9 (0.1) |
| Tofacitinib, n (%) | 30 (0.3) |
| Baricitinib, n (%) | 28 (0.2) |
| Infliximab biosimilar, n (%) | 26 (0.2) |
| Etanercept biosimilar, n (%) | 59 (0.5) |
| NSAIDs, n (%) | 6339 (53.4) |
| Gastric antacid, n (%) | 7550 (63.6) |
| Folic acid, n (%) | 5206 (43.9) |
| Medication for osteoporosis, n (%) | 3937 (33.2) |
| Medical history |  |
| Angina pectoris or Myocardial infarction, n (%) | 617 (5.2) |
| Heart failure, n (%) | 279 (2.4) |
| Stroke, n (%) | 285 (2.4) |
| High blood pressure, n (%) | 3216 (27.1) |
| Interstitial pneumonia, n (%) | 735 (6.2) |
| Gastric ulcer, n (%) | 1436 (12.1) |
| Diabetes mellitus, n (%) | 872 (7.3) |
| Cancer, n (%) | 853 (7.2) |
| Depression, n (%) | 512 (4.3) |
| Amyloidosis, n (%) | 8 (0.1) |
| Skin ulcer, n (%) | 83 (0.7) |

BMI, body mass index; CDAI, Clinical Disease Activity Index; DAS, Disease Activity Score; ESR, Erythrocyte Sedimentation Rate; J-HAQ, Japanese Health Assessment Questionnaire; NSAIDs, Non-Steroidal Anti-Inflammatory Drugs; SD, standard deviation.

**Table S3. DAS28-ESR calculation responses from Llama2 and Meditron models**

Answer of Llama2

The DAS28-ESR is a measure of disease activity in patients with rheumatoid arthritis (RA). It is calculated using a combination of clinical and laboratory variables. The variables required for the DAS28-ESR calculation are:

Swollen joint count (SJC): The number of joints that are swollen and painful due to inflammation.

Tender joint count (TJC): The number of joints that are tender to the touch due to inflammation.

Patient global assessment (PGA): The patient's subjective assessment of their overall disease activity, measured on a visual analog scale (VAS) from 0 (no activity) to 100 (maximum activity).

Erythrocyte sedimentation rate (ESR): A measure of inflammation in the body, measured in millimeters per hour (mm/h).

The calculation formula for the DAS28-ESR is as follows:

DAS28-ESR = 0.56 ¥√(SJC + TJC) + 0.28 ¥ PGA + 0.11 ¥ ESR

The DAS28-ESR score ranges from 0 to 100, with higher scores indicating greater disease activity.

Answer of Meditron

\# Introduction

The Disease Activity Score (DAS) is a composite index that combines information from the number of swollen joints, the number of tender joints, the patient's global assessment of disease activity, and the erythrocyte sedimentation rate (ESR) or C-reactive protein (CRP) level. The DAS28-ESR is a version of the DAS that uses the ESR. The DAS28-ESR is used to assess the disease activity of patients with rheumatoid arthritis (RA). The DAS28-ESR is calculated using the following formula:

```
## Das28-esr = 0.56 × √(tender28) + 0.28 × √(swollen28) + 0.70 × ln(esr) + 0.014 × global
## Tender28
```

The number of tender joints out of 28 joints.

\## Swollen28

The number of swollen joints out of 28 joints.

\## ESR

The erythrocyte sedimentation rate (mm/h).

\## Global

The patient's global assessment of disease activity (0-100 mm).

\## Interpretation

The DAS28-ESR is interpreted as follows:

\## Remission

DAS28-ESR < 2.6.

\## Low disease activity

DAS28-ESR ≥ 2.6 and < 3.2.

\## Moderate disease activity

DAS28-ESR ≥ 3.2 and < 5.1.

\## High disease activity

DAS28-ESR ≥ 5.1.

**Table S4**. **Comparison of AUC between Llama2 and Meditron models**

|  | Llama2  without fine-tuning | Meditron  without fine-tuning | *P*† |
| --- | --- | --- | --- |
| DAS28-ESR >5.1 | 0.76 | 0.82 | < 0.001 |
| DAS28 ESR <2.6 | 0.80 | 0.83 | < 0.0001 |
| DAS28 CRP >4.1 | 0.78 | 0.79 | 0.55 |
| DAS28 CRP <2.3 | 0.76 | 0.77 | 0.028 |
| CDAI >22 | 0.81 | 0.76 | < 0.0001 |
| CDAI ≤2.8 | 0.76 | 0.72 | < 0.0001 |
| J-HAQ >2.5 | 0.90 | 0.96 | 0.037 |
| J-HAQ <0.5 | 0.80 | 0.92 | < 0.0001 |

†: DeLong's test

CDAI, Clinical Disease Activity Index; CRP, C-Reactive Protein; DAS, Disease Activity Score; ESR, Erythrocyte Sedimentation Rate; J-HAQ, Japanese Health Assessment Questionnaire.

**Table S5**. **Comparison of AUC between fine-tuned and non-fine-tuned Llama2 and Meditron models**

|  | fine-tuning (-) | fine-tuning (+) | *P*† |
| --- | --- | --- | --- |
| Llama2  DAS28 ESR >5.1 | 0.76 | 0.83 | < 0.001 |
| DAS28 ESR <2.6 | 0.80 | 0.85 | < 0.0001 |
| DAS28 CRP >4.1 | 0.78 | 0.85 | < 0.0001 |
| DAS28 CRP <2.3 | 0.76 | 0.80 | < 0.0001 |
| CDAI >22 | 0.81 | 0.84 | 0.35 |
| CDAI ≤2.8 | 0.76 | 0.78 | 0.042 |
| J-HAQ >2.5 | 0.90 | 0.96 | < 0.0001 |
| J-HAQ <0.5 | 0.80 | 0.92 | < 0.0001 |
| Meditron  DAS28 ESR >5.1 | 0.82 | 0.83 | 0.27 |
| DAS28 ESR <2.6 | 0.83 | 0.85 | < 0.0001 |
| DAS28 CRP >4.1 | 0.79 | 0.82 | 0.0015 |
| DAS28 CRP <2.3 | 0.77 | 0.81 | < 0.0001 |
| CDAI >22 | 0.76 | 0.81 | 0.061 |
| CDAI ≤2.8 | 0.72 | 0.79 | < 0.0001 |
| J-HAQ >2.5 | 0.96 | 0.96 | 0.13 |
| J-HAQ <0.5 | 0.92 | 0.92 | 0.091 |

†: DeLong's test

CDAI, Clinical Disease Activity Index; CRP, C-Reactive Protein; DAS, Disease Activity Score; ESR, Erythrocyte Sedimentation Rate; J-HAQ, Japanese Health Assessment Questionnaire.

**Table S6**. **Comparison of AUC between linear regression and Fine-tuned LLM models**

|  | Linear regression | LLM | *P*† |
| --- | --- | --- | --- |
| Llama2  DAS28 ESR >5.1 | 0.84 | 0.83 | 0.44 |
| DAS28 ESR <2.6 | 0.83 | 0.85 | 0.0042 |
| DAS28 CRP >4.1 | 0.78 | 0.85 | < 0.0001 |
| DAS28 CRP <2.3 | 0.78 | 0.80 | 0.020 |
| CDAI >22 | 0.85 | 0.84 | 0.58 |
| CDAI ≤2.8 | 0.80 | 0.78 | 0.0082 |
| J-HAQ >2.5 | 0.80 | 0.96 | < 0.0001 |
| J-HAQ <0.5 | 0.80 | 0.92 | < 0.0001 |
| Meditron  DAS28 ESR >5.1 | 0.84 | 0.83 | 0.54 |
| DAS28 ESR <2.6 | 0.83 | 0.85 | 0.012 |
| DAS28 CRP >4.1 | 0.78 | 0.82 | 0.0087 |
| DAS28 CRP <2.3 | 0.78 | 0.81 | 0.0010 |
| CDAI >22 | 0.85 | 0.81 | 0.14 |
| CDAI ≤2.8 | 0.80 | 0.79 | 0.058 |
| J-HAQ >2.5 | 0.80 | 0.96 | < 0.0001 |
| J-HAQ <0.5 | 0.80 | 0.92 | < 0.0001 |

†: DeLong's test

CDAI, Clinical Disease Activity Index; CRP, C-Reactive Protein; DAS, Disease Activity Score; ESR, Erythrocyte Sedimentation Rate; J-HAQ, Japanese Health Assessment Questionnaire.

**Table S7. DAS28-ESR calculation responses from fine-tuning Meditron models**

Answer of fine-tuned Meditron

The DAS28-ESR is calculated as follows:

DAS28-ESR = 0.56*sqrt(TJC28) + 0.28*sqrt(SJC28) + 0.70*ln(ESR) + 0.014*GH

TJC28 = tender joint count of 28 joints

SJC28 = swollen joint count of 28 joints

ESR = erythrocyte sedimentation rate

GH = general health (0-100 mm visual analogue scale)

The DAS28-ESR is interpreted as follows:

DAS28-ESR < 2.6 = remission

2.6 ≤ DAS28-ESR < 3.2 = low disease activity

3.2 ≤ DAS28-ESR < 5.1 = moderate disease activity

DAS28-ESR ≥ 5.1 = high disease activity
